# Novel COVID-19 phenotype definitions reveal phenotypically distinct patterns of genetic association and protective effects

**DOI:** 10.1101/2021.01.24.21250324

**Authors:** Genevieve H.L. Roberts, Raghavendran Partha, Brooke Rhead, Spencer C. Knight, Danny S. Park, Marie V. Coignet, Miao Zhang, Nathan Berkowitz, David A. Turrisini, Michael Gaddis, Shannon R. McCurdy, Milos Pavlovic, Luong Ruiz, AncestryDNA Science Team, Asher K. Haug Baltzell, Harendra Guturu, Ahna R. Girshick, Kristin A. Rand, Eurie L. Hong, Catherine A. Ball

## Abstract

Multiple large COVID-19 genome-wide association studies (GWAS) have identified reproducible genetic associations indicating that some infection susceptibility and severity risk is heritable.^1-5^ Most of these studies ascertained COVID-19 cases in medical clinics and hospitals, which can lead to an overrepresentation of cases with severe outcomes, such as hospitalization, intensive care unit admission, or ventilation. Here, we demonstrate the utility and validity of deep phenotyping with self-reported outcomes in a population with a large proportion of mild and subclinical cases. Using these data, we defined eight different phenotypes related to COVID-19 outcomes: four that align with previously studied COVID-19 definitions and four novel definitions that focus on susceptibility given exposure, mild clinical manifestations, and an aggregate score of symptom severity. We assessed replication of 13 previously identified COVID-19 genetic associations with all eight phenotypes and found distinct patterns of association, most notably related to the chr3/*SLC6A20/LZTFL1* and chr9/*ABO* regions. We then performed a discovery GWAS, which suggested some novel phenotypes may better capture protective associations and also identified a novel association in chr11/*GALNT18* that reproduced in two fully independent populations.

## MAIN TEXT

To perform genetic studies of COVID-19, we conducted a comprehensive, 50+ question survey of AncestryDNA customers that assessed exposure, risk factors, symptomatology, and demographic information **(Supplementary Figure 1; Supplementary Table 1)**. We collected over 700,000 COVID-19 survey responses between April and August 2020 and used them to develop an expanded repertoire of phenotypes to investigate. In total, we defined eight COVID-19 phenotypes, summarized in **Table 1**. Four phenotypes were intended to mirror susceptibility or severity phenotype definitions from other large COVID-19 GWAS^2,3^ and four are novel. We hypothesized that novel phenotype definitions focusing on mild outcomes or absence of infection despite a strong exposure may be better suited to detecting protective genetic associations than traditional phenotypes.

**Table 1.** Summary of Eight Phenotype Definitions.

| Phenotype Code<sup>1</sup> | Case Description<sup>2</sup> | Control Description<sup>2</sup> | Novelty | Type | Goal |
| --- | --- | --- | --- | --- | --- |
| *Positive/Negative* | **COVID-19(+)** | COVID-19(-) | Traditional | Susceptibility | Reproduce other studies |
| *Positive/Unscreened* | **COVID-19(+)** | Unscreened, but not known to be COVID-19(+) | Traditional | Susceptibility | Reproduce other studies |
| *Hospitalized/Not_Hospitalized* | **COVID-19(+) and hospitalized** | COVID-19+ and not hospitalized | Traditional | Severity | Reproduce other studies |
| *Hospitalized/Unscreened* | **COVID-19(+) and hospitalized** | Unscreened, but not known to be COVID-19(+) | Traditional | Severity | Reproduce other studies |
| *Exposed_Positive/Exposed_Negative* | COVID-19(+) and had a housemate with a confirmed COVID-19 diagnosis | **COVID-19(-) and had a housemate with a confirmed COVID-19 diagnosis** | Novel | Susceptibility | Study genetic susceptibility in individuals thought to have a strong exposure event |
| *Unscreened/Exposed_Negative* | Unscreened, but not known to be COVID-19(+) | **COVID-19(-) and had a housemate with a confirmed COVID-19 diagnosis** | Novel | Susceptibility | Study genetic protection from infection in individuals thought to have a strong exposure event |
| *Symptomatic/Paucisymptomatic* | COVID-19(+) and symptomatic | **COVID-19(+) and asymptomatic or paucisymptomatic** | Novel | Severity | Study genetic protection from severe outcomes if infected |
| *Continuous_Severity_Score*<sup>3</sup> | COVID-19(+) score that combines nine different measures of COVID-19 symptom severity. Higher scores correspond to more severe outcomes. |  | Novel | Severity | Study genetic variants associated with both severe and mild outcomes simultaneously |

1. Nomenclature for phenotype codes: Case\_definition/Control\_definition
2. Case or Control Descriptions in bold represent the minority group
3. The Continuous\_Severity\_Score phenotype is continuous, and thus there are no cases and controls. Instead, a score is computed for each person.

**Figure 1.**
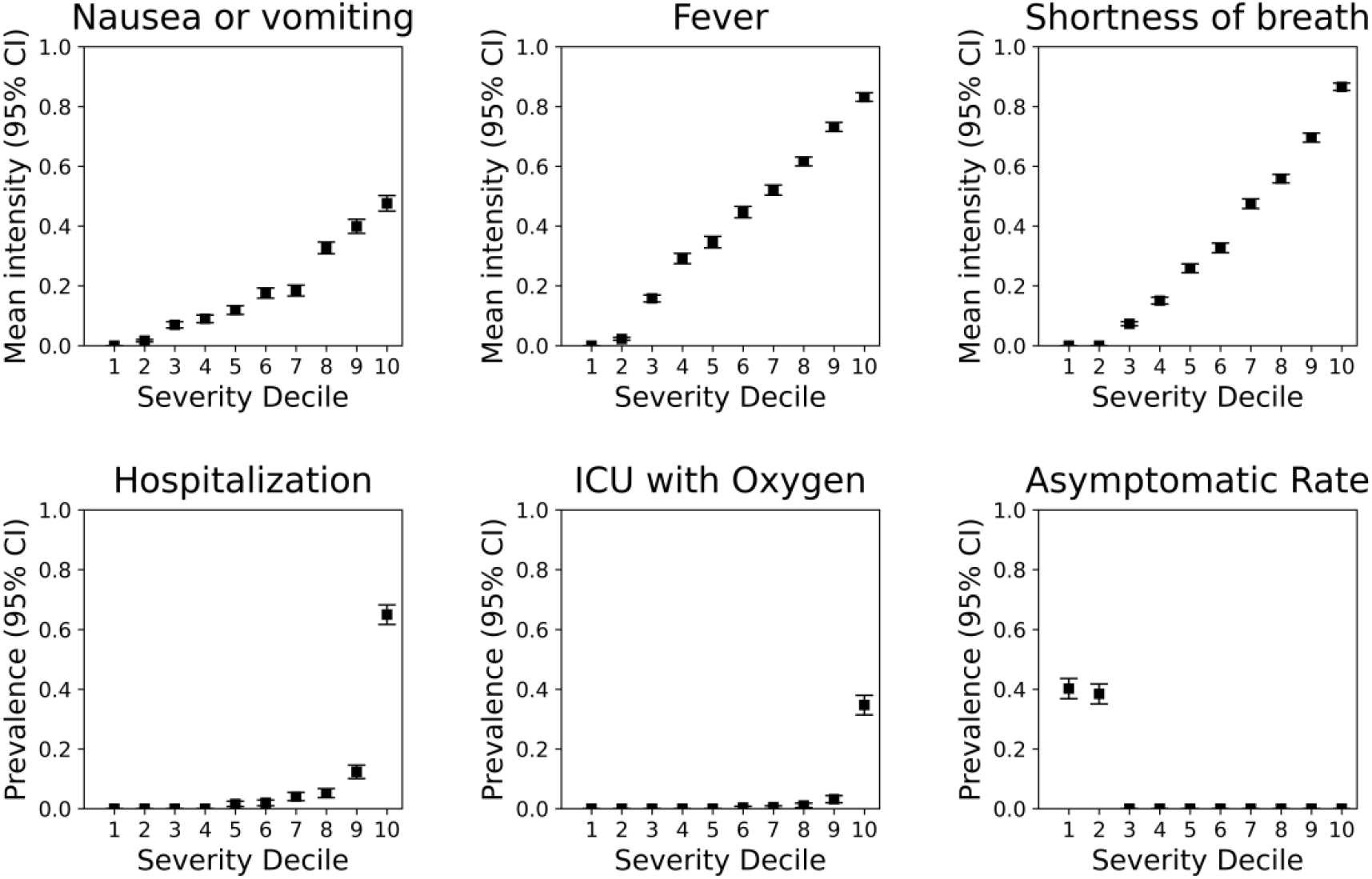
COVID-19 Continuous Severity Score Captures Multiple Aspects of Symptom Severity Among COVID-19(+) Individuals. The continuous severity score was derived from the first principal component across nine survey fields related to COVID-19 clinical outcomes, including three symptoms, hospitalization, ICU admittance, and other severe complications due to COVID-19 illness (see Methods). Plots reflect mean symptom severity (top three panels) or prevalence (bottom three panels) for several fields as a function of ascending severity decile. Symptom information was encoded as follows: 0=None, 0.2=Very Mild, 0.4=Mild, 0.6=Moderate, 0.8=Severe, and 1.0=Very Severe. A paucisymptomatic case corresponds to reporting symptoms of mild intensity or less. Squares represent the estimate and vertical lines represent the 95% confidence intervals for each estimate.

Susceptibility to infection is difficult to measure because contracting the virus depends on exposure. We therefore designed two novel susceptibility phenotypes that focus on respondents with a known, strong exposure to the virus—those who had “household exposure.” The positivity rate among respondents that reported a housemate with confirmed COVID-19 was approximately 65%, the highest positivity rate for any exposure we assessed. The *Exposed_Positive/Exposed_Negative* phenotype compared those with a household exposure that tested positive to those with a household exposure that tested negative, and *Unscreened/Exposed_Negative* focused on protection from infection by comparing those with a household exposure that tested negative to a large sample of unscreened controls. We also defined two novel severity phenotypes: *Symptomatic/Paucisymptomatic*, which compares cases with symptomatic infections to those with very mild or asymptomatic infections, and *Continuous_Severity_Score* which unifies asymptomatic and severely ill COVID-19 patients. The *Continuous_Severity_Score* aggregates responses from nine survey fields. Lower scores correspond to lower symptom severity, while higher correspond to increased symptom severity and elevated hospitalization rates (**Figure 1**). Sample sizes for each phenotype are presented in **Supplementary Table 2**. For all eight phenotypes, cases corresponded to higher risk of susceptibility or severity so that all positive SNP effect estimates 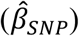 can be interpreted as “risk” and all negative 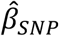 can be interpreted as “protective.”

Our first goal was to explore how known COVID-19 risk loci associate with the different phenotype definitions. To accomplish this, we identified 13 independent SNPs (*r*^*2*^<0.05) that achieved genome-wide significance in at least one of two recent, large, COVID-19 meta-analyses: the October 2020 data release from the COVID-19 Host Genetics Initiative (HGI) or Horowitz *et al*. **(Supplementary Table 3)**. We assessed association of these 13 SNPs with all eight phenotypes in a trans-ancestry meta-analysis of European (EUR), Admixed Amerindian (LAT), and Admixed African-European (AA) cohorts **(Supplementary Figure 2)**. We considered a trans-ancestry *P*-value of <0.05 and consistent direction of 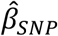 with the prior study evidence of replication **(Supplementary Table 4)**. We note that a small percentage of research participants in our study overlaps prior studies, quantified in **Supplementary Figure 3**.

**Figure 2:**
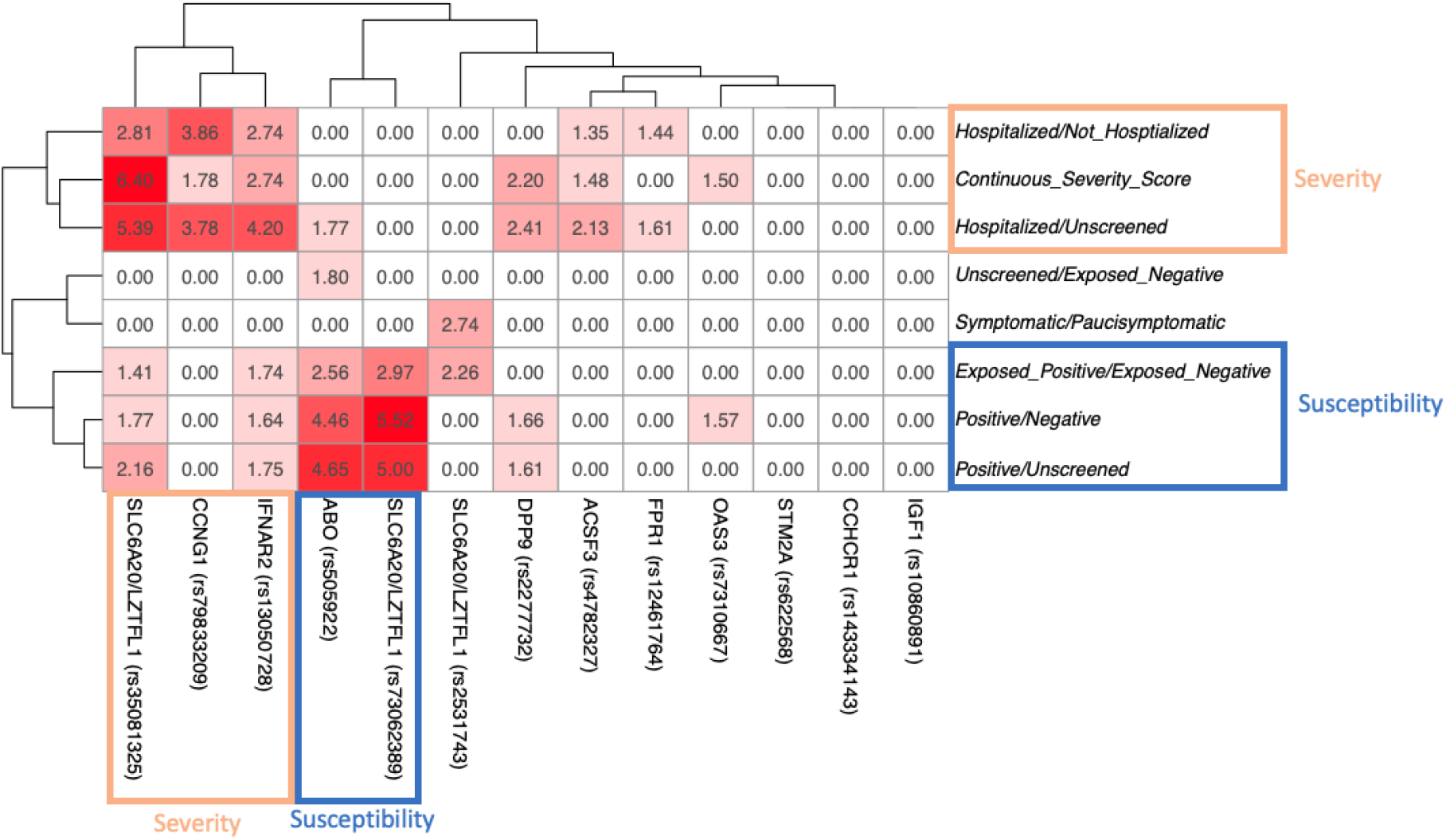
Heatmap of replication at 13 lead SNPs identified by previous studies. Each pairwise block represents the trans-ancestry meta-analysis −log_10_(*P*-value) for the association between one of the eight phenotypes we examined, and one of 13 loci previously identified by Horowitz *et al*. and/or HGI. Red blocks denote replication, with darker shades of red corresponding to lower trans-ancestry *P*-values in our analysis, and white blocks representing no association. All associations with trans-ancestry *P*>0.05 or with inconsistent directions of effect relative to the previous study were forced to have −log_10_(*P*-value)=0. SNP and phenotype labels were ordered by hierarchical clustering, with corresponding dendrograms shown on the top and left of the figure. Orange rectangles annotate phenotypes or loci that appear to associate more strongly with severity whereas blue rectangles annotate phenotypes or loci that appear to associate more strongly with susceptibility. Extended summary statistics for all associations in all studies are available in **Supplementary Table 4**.

**Figure 3.**
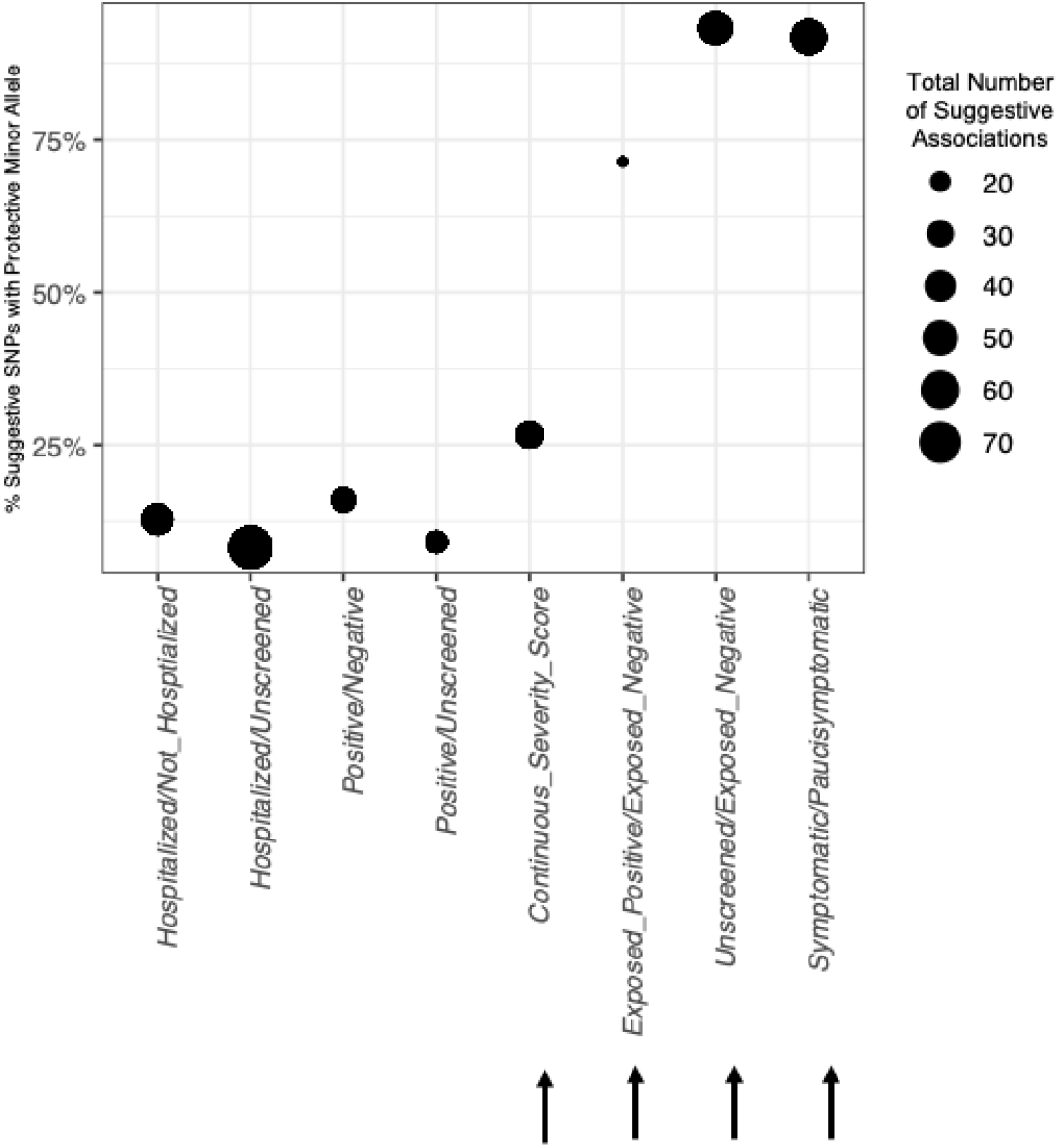
Novel Phenotypes Detect More Associations with a Protective Minor Allele. The size of each point represents the total number of novel, suggestive SNPs (discovery EUR *P*<1×10^−5^) for each of the eight phenotypes. The y-axis position of each point shows the percentage of suggestively associated SNPs for which the discovery EUR minor allele was in the protective direction of effect. Arrows show the four novel phenotype definitions.

Replication results are visualized in **Figure 2**. Ten of 13 SNPs replicated in at least one of our phenotypes. This result demonstrates that our phenotypes, which are based on self-reported outcomes, strongly recapitulate the same associations previously found by clinical phenotyping. Hierarchical clustering of the replication *P*-values revealed two unique clusters of phenotype-locus pairs: three *severity* phenotypes produced a similar pattern of replication (*Hospitalized/Not_Hosptialized, Hospitalized/Unscreened, Continuous_Severity_Score*) and three *susceptibility* phenotypes produced a similar pattern of replication (*Positive/Negative, Positive/Unscreened, Exposed_Positive/Exposed_Negative*). Phenotypes in these clusters are likely capturing similar genetic associations; however, the strength of associations differ, suggesting that some phenotype definitions are more powerful than others. The two remaining novel phenotypes (*Symptomatic/Paucisymptomatic, Unscreened/Exposed_Negative*) replicated the 13 SNPs poorly and may capture different genetic associations, warranting further investigation.

The patterns of locus replication are of special interest, particularly in chr3 and chr9 regions. There are three independent signals in a 52Kb region on chr3 near a cluster of immune genes including *LZTFL1* and *SLC6A20*. The main HGI *severity* study (“ANA_B2”) identified rs35081325, which is strongly associated with the *severity* cluster of phenotypes. Thus, rs35081325 appears to consistently associate with increased risk of infection severity. By contrast, rs73062389 was identified in the main HGI *susceptibility* study (“ANA_C2”) and is strongly associated with the *susceptibility* cluster of phenotypes. Furthermore, rs73062389 is not associated with *any* of our severity cluster phenotypes and thus seems to specifically confer increased *susceptibility* risk. Finally, rs2531743, a novel signal recently discovered by Horowitz *et al*. in an analysis of severity, associated with only two phenotypes in our study: *Symptomatic/Paucisymptomatic* and *Exposed_Positive/Exposed_Negative*. Unlike the other chr3 signals, the minor allele of rs2531743 is associated in the protective direction of effect. Thus, all three signals in this region associate with a totally distinct set of phenotypes.

Associations near *ABO*, the gene that determines blood type, have also been observed in multiple COVID-19 GWAS—somewhat inconsistently with severity phenotypes and more consistently with susceptibility phenotypes **(Supplementary Table 5)**. The lead *ABO* SNP, rs505922, replicated in all four susceptibility phenotypes plus one severity phenotype. The only severity phenotype associated with the *ABO* SNP was *Hospitalized/Unscreened*, which utilized a large number of unscreened controls. We speculate that unscreened controls induce susceptibility associations because hospitalized cases *must* be susceptible to infection, but the unscreened control group *may or may not* be susceptible, and thus this phenotype simultaneously captures aspects of both susceptibility and severity.

Our second goal was to discover novel phenotype-locus associations; we therefore conducted a discovery GWAS for all eight phenotypes. Due to the novelty of the phenotypes and the difficulty in obtaining a truly independent COVID-19 replication cohort, we opted to conduct the discovery GWAS in the same EUR cohort used in the above trans-ancestry meta-analysis, and dedicate a smaller, fully independent EUR cohort, the LAT cohort, and the AA cohort to determining whether any newly identified phenotype-locus associations reproduce.

No phenotype-locus association pairs surpassed a conservative Bonferroni-corrected significance threshold of discovery *P*<6.25×10^−9^, but we examined associations that reached a suggestive significance threshold of *P*<1×10^−5^ to look for trends. In total, we identified 297 suggestive phenotype-locus association pairs (**Supplementary Table 6**). Strikingly, minor alleles suggestively associated with three novel phenotypes (*Exposed_Positive/Exposed_Negative, Unscreened/Exposed_Negative, Symptomatic/Paucisymptomatic*) were *nearly always* associated with a *protective* direction of effect, whereas for all previously studied phenotypes, the minor allele was *nearly always* associated in the *risk* direction **(Figure 3)**. This finding supports our hypothesis that the novel phenotype definitions that focus on mild outcomes or absence of infection despite a strong exposure may be better suited to detecting protective genetic associations than traditional phenotypes.

Overall, we observed low rates of replication among the 297 phenotype-locus association pairs (mean replication rate=3.7%; **Supplementary Table 7**) and low correlation of 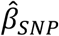 across the three independent populations (mean 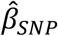 Pearson *R*=-0.23; **Supplementary Figure 4; Supplementary Table 7**). This result suggests that the independent replication cohorts had insufficient power or that many of the suggestive phenotype-locus pairs simply represent false-positive associations. Interestingly, however, two novel phenotypes generally had positive 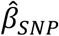 correlations across independent populations: *Continuous_Severity_Score* and *Exposed_Positive/Exposed_Negative* **(Supplementary Figure 4b-c)**, suggesting that these phenotypes might yield reproducible associations as replication cohort sample sizes grow larger. There were also 15 phenotype-locus association pairs that reproduced in one independent population, and one that replicated in two independent populations (**Supplementary Table 8**). The phenotype-locus association that replicated in two fully independent populations was *Hospitalized/Not_Hosptialized* with rs55673936 (**Figure 4)**. This SNP is an intronic variant on chr11 in the gene *GALNT18*. Interestingly, another SNP within *GALNT18* was previously reported as associated with an increased response to Tocilizumab^6^, an IL-6 blocking monoclonal antibody that has been tested in multiple clinical trials for treatment of COVID-19, albeit with mixed preliminary success.^7-9^ Nonetheless, this novel association with *GALNT18* complements findings by other genetic studies that point to modulation of the IL-6 pathway as a potential strategy to ameliorate severe COVID-19 in some people.^10^

**Figure 4.**
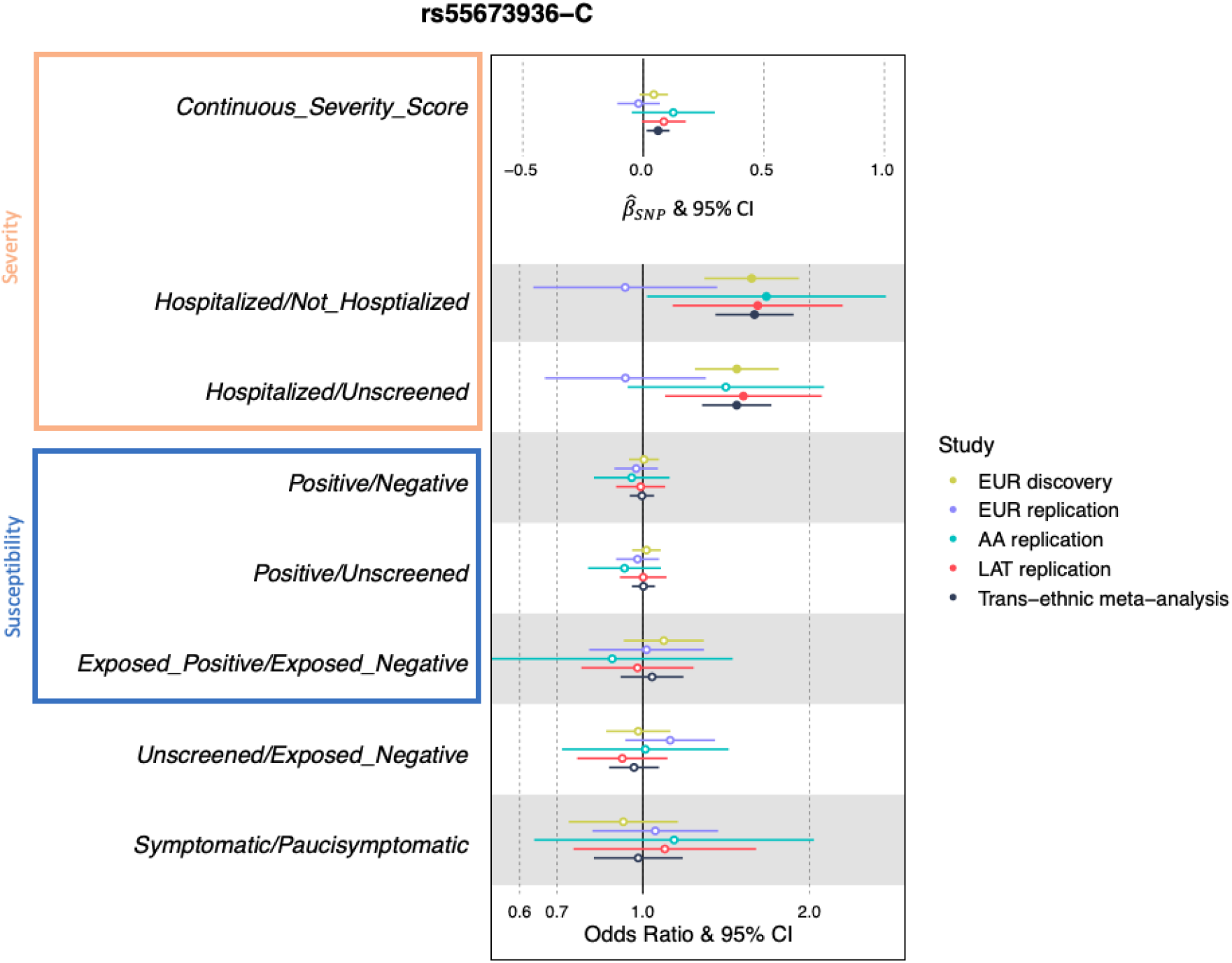
Forest Plot of Novel Association with *GALNT18* intronic SNP, rs55673936-C, with the eight phenotypes. Circles indicate effect estimates and horizontal lines represent 95% confidence intervals. *Continuous_Severity_Score* was the only continuous phenotype and therefore the reported effect estimate is the 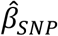, which can be interpreted as severity score standard deviations from the mean per each copy of the “C” minor allele. For all other phenotypes, per-allele odds ratios are reported. Filled circles indicate *P*<0.05. The orange rectangle annotates phenotypes in the severity cluster and the blue rectangle annotates the susceptibility cluster, with clusters defined in **Figure 2**.

In summary, we explored genetic association with eight different COVID-19 phenotype definitions, four of which have not yet been explored. We find that 10 of 13 previously identified COVID-19 genetic signals associate with at least one of the eight phenotype definitions. This strong replication of loci identified by clinically ascertained studies confirms that phenotyping based on well-designed self-report studies is valid. Some of these replicated genetic signals clearly associate more with severity phenotypes and others associate more with susceptibility phenotypes, suggesting that heterogeneity in ascertainment and different case/control definitions likely underlies inconsistent associations, for instance *ABO*. Our findings also show that all three previously identified signals in the chr3 *LZTFL1*/*SLC6A20* region associate with a different set of phenotypes, suggesting that variation in this region modulates multiple aspects of COVID-19 susceptibility and severity and thus is extremely important. In our discovery analysis, we identified a novel association with rs55673936, a *GALNT18* intron variant that reproduced in multiple independent populations. Whereas other groups with primary ascertainment at medical clinics are better equipped to study severe outcomes, our self-reported dataset allows a complementary analysis of more granular phenotypes in a population enriched for mild outcomes. We find promising evidence that exploring new phenotypes in this unique population will yield novel genetic associations, particularly those that confer protection against the novel coronavirus.

## ONLINE METHODS

### Ethics statement

All data for this research project were from subjects who provided prior informed consent to participate in AncestryDNA’s Human Diversity Project, as reviewed and approved by our external institutional review board, Advarra (formerly Quorum). All data were de-identified prior to use.

### Study population

Self-reported COVID-19 outcomes were collected through the Personal Discoveries Project®, a survey platform available to AncestryDNA customers via the web and mobile applications. The COVID-19 survey ranged from 39-71 questions, depending on the initial COVID-19 test result reported. **Supplementary Figure 1** describes the flow of the topics assessed in each section of the survey. Analyses presented here were performed with data collected between April 22-August 3, 2020.

To participate in the COVID-19 survey, participants must meet the following criteria: they must be 18 years of age or older, a resident of the United States, be an existing AncestryDNA customer who has consented to participate in research and be able to complete a short survey. The survey is designed to assess self-reported COVID-19 positivity and severity, as well as susceptibility and known risk factors including community exposure and known contacts with individuals diagnosed with COVID-19.

### Binary Phenotype Definitions

In total, we assessed eight phenotypes, which are summarized in **Table 1**. Key definitions include testing positive or negative, hospitalization, asymptomatic cases, and housemate exposure. COVID-19 positivity or negativity was assessed by the question “Have you been swab tested for COVID-19, commonly referred to as coronavirus?”. Hospitalization due to COVID-19 illness was used as one binary measure of severity, and was assessed with the question, “Were you hospitalized due to these symptoms?”. Asymptomatic cases were defined as those that were positive for COVID-19 and either answered “No” do the question “Did you experience symptoms as a result of your condition?” or answered either “None”, “Very mild”, or “Mild” to all 15 questions related to symptom severity. High exposure to COVID-19 was assessed through having a positive housemate, assessed by the question, “Has someone in your household tested positive for COVID-19?”.

### Continuous Severity Phenotype Creation

A continuous severity score was derived by computing the first principal component across nine survey fields related to COVID-19 clinical outcomes. Six of the nine questions were binary: hospitalization, intensive care unit (ICU) admittance with oxygen, ICU admittance with ventilation, septic shock, respiratory failure, and organ failure due to COVID-19. Binary responses were encoded as 0 for “No” and 1 for “Yes”. Three symptom questions related to shortness of breath, fever, and nausea/vomiting symptoms were encoded as a unit-scaled variable based on the following mapping: 0=“None”, 0.2=“Very mild”, 0.4=“Mild”, 0.6=“Moderate”, 0.8=“Severe”, and 1.0=“Very Severe”. The three symptoms were chosen based on prior literature indicating their positive association with COVID-19 hospitalization.^11^ The following assumptions were made to so that a score could be calculated for most participants who reported a positive COVID-19 test:

- Participants who responded “No” to the question “Did you experience symptoms as a result of your condition?” were not presented with additional questions regarding symptomatology or hospitalization and thus were encoded as 0 for all individual symptoms (shortness-of-breath, fever, nausea/vomiting), hospitalization, ICU admittance, and severe complications due to COVID-19 illness.
- Participants who responded “No” to the question “Were you hospitalized due to these symptoms?” were not presented any further questions regarding hospitalization and thus were encoded as 0 for ICU admittance and supplemental oxygen.
- Participants who declined to answer a question about complications due to COVID-19 illness such as septic shock, respiratory failure, and organ failure were encoded as 0 for those complications (<2% of all participants for whom continuous severity was scored).

### Genotyping

Genotyping and quality control procedures have been previously described elsewhere.^12^ Briefly, customer genotype data for this study were generated using an Illumina genotyping array and processed either with Illumina or with Quest/Athena Diagnostics. To ensure quality of each dataset, a sample passes a number of quality control (QC) checks, which includes identifying duplicate samples, removing individuals with a per-sample call rate <98%, and identifying discrepancies between reported sex and genetically inferred sex. Samples that pass all quality-control tests proceed to the analysis pipeline; samples that fail one or more tests must be recollected or manually cleared for analysis by lab technicians. Array markers with per-variant call rate <0.98 and array markers that had overall allele frequency differences of >0.10 between any two array versions were additionally removed prior to downstream analyses.

### Defining ancestry cohorts

We defined three separate ancestry cohorts: a European ancestry group (EUR), an Admixed Amerindian ancestry (LAT), and an Admixed African ancestry group (AA) **(Supplementary Figure 2)**. We assigned COVID-19 survey respondents to one of these ancestry groups with a proprietary algorithm that estimates continental admixture proportions. Briefly, this algorithm uses a hidden Markov model to estimate unphased diploid ancestry across the genome by comparing haplotype structure to a reference panel. The reference panel consists of a combination of AncestryDNA customers and publicly available datasets and is designed to reflect global diversity. From our total cohort of 736,723 individuals who participated in the COVID-19 survey as of August 3, 2020, 537,512 (73%) individuals were designated to the EUR group, 22,464 (3%) to the AA group, and 47,301 (6%) to the (LAT) group, and the remainder were not assigned to any ancestry group **(Supplementary Table 1)**.

### Removal of related individuals

AncestryDNA’s identity-by-descent inference algorithm^13^ was used to estimate the relationship between pairs of individuals. Pairs with estimated separation of fewer than four meioses were considered close relatives. For all close relative pairs, one individual was randomly selected for exclusion from our study. In total, we excluded ∼8% (60,379) individuals from analysis due to relatedness.

### Calculation of principal components (PCs)

For each population described above, genetic PCs were calculated to include in the association studies to control residual population structure and were computed using FlashPCA 2.0.^14^ Input genotypes were linkage disequilibrium (LD)-pruned using PLINK 1.9 command _--indep-pairwise 100 5 0.2 -- maf 0.05 --geno 0.001_.

### Imputation

Samples were imputed to the Haplotype Reference Consortium (HRC) reference panel version 1.1, which consists of 27,165 total individuals and 36 million variants. The HRC reference panel does not include indels; consequently, indels are not present in the results of our analyses. We determined best-guess haplotypes with Eagle^15^ version 2.4.1 and performed imputation with Minimac4 version 1.0.1. We used 1,077,214 unique variants as input and 8,187,660 imputed variants were retained in the final data set. For these variants, we conservatively restricted our analyses to variants with minor allele frequency (MAF)>0.01 and Minimac4 R^2^>0.30 using imputed dosages for all variants regardless of whether they were originally genotyped.

### Discovery GWAS

Discovery GWAS were conducted in EUR ancestry only. For discovery, we conducted sex-stratified GWAS and meta analyze the results via inverse-variance weighting implemented with METAL^16^(version released 25 March 2011). For each phenotype, a GWAS assuming an additive genetic model was implemented with PLINK2.0. Imputed genotype dosage value was the primary predictor. The following were included as fixed-effect covariates: PCs 1-25 (described above), array platform, orthogonal age, and orthogonal age^2^. Orthogonal polynomials were used to eliminate collinearity between age and age^2^ and were calculated in R version 3.6.0 with base function poly(age, degree=2). We additionally used PLINK2.0 to remove variants with Minimac4 imputation quality *R*^*2*^<0.3 or with MAF<0.01. The following PLINK2.0 flags were used for each analysis:

> --vcf [input imputed VCF] dosage=DS
>
> --psam [file that provides sex information]
>
> --covar [covariates file]
>
> --covar-name PC1, PC2, PC3, PC4, PC5, PC6, PC7, PC8, PC9, PC10, PC11, PC12, PC13, PC15, PC15, PC16, PC17, PC18, PC19, PC20, PC21, PC22, PC23, PC24, PC25, orthogonal_age, orthogonal_age2, platform
>
> --covar-variance-standardize
>
> --extract-if-info R2 >= 0.3
>
> --freq
>
> --glm
>
> --keep [list of unrelated Europeans]
>
> --keep-females OR keep-males
>
> --maf 0.01
>
> --pheno [1 of 8 phenotype files]
>
> --pheno-name [phenotype column name]

Unless otherwise noted, all EUR discovery variant effect estimates are adjusted for the 28 covariates described above. To establish significance, we implemented a stringent, Bonferroni-corrected significance threshold by dividing the typical genome-wide significance threshold in Europeans of *P*<5×10^−8^ by the eight phenotypes, which results in *P*<6.25×10^−9^. Suggestive significance followed the definition used by the HGI consortium of *P*<1×10^−5^.

### Independent Replication GWAS

We used the AA, LAT, and a smaller, fully independent EUR cohort to replicate our findings. We began reserving respondents for the replication EUR cohort at the conclusion of our previous study^12^ on May 28, 2020, and thus the replication EUR cohort is not representative of the full period of survey collection. The AA and LAT cohorts were steadily collected throughout the entire collection period that spanned April 22, 2020 to August 3, 2020. We conducted separate GWAS for each of these three replication cohorts and for each of the eight phenotypes. The same association procedure that was used for the discovery study was applied for replication cohorts, except sample sizes for these cohorts were smaller (**Supplementary Table 2**), and thus a single GWAS was conducted for males and females together with genetic sex included as a covariate.

### Trans-Ancestry Meta-Analysis

For each phenotype, we additionally performed a trans-ancestry meta-analysis of the discovery EUR cohort, AA, and LAT summary statistics, again using fixed-effect inverse-variance weighting implemented in METAL. The replication EUR cohort was not included in the trans-ancestry meta-analysis. These summary statistics were used to assess replication of the 13 loci defined in the next section.

### Replication of 13 Independent SNPs from Previous Studies

We manually curated a list of 13 independent SNPs that represent lead loci identified by either HGI or Horowitz *et al*. Eight of the 13 SNPs were lead SNPs in HGI’s most recent data release (October 2020; without 23andMe data included). These eight SNPs were the most-associated marker at any locus achieving *P*<5×10^−8^ in the Hospitalization vs. Population (“ANA_B2”) or COVID-19(+) vs. Population (“ANA_C2”). The remaining five SNPs were selected from Figure 1 of a recent trans-ancestry meta-analysis.^2^ We note that a subset of AncestryDNA survey respondents overlap those included in the large meta analyses conducted by HGI and Horowitz *et al*. and thus replication in our study is not completely independent **(Supplementary Figure 3)**. All 13 SNPs in the final list are independent of one another (*r*^*2*^<0.05) and represent 11 positionally distinct loci (>500Kb apart). One of the 11 loci encompasses three independent SNPs that span a 52Kb region near *SLC6A20/LZTFL1* on chr3. For these 13 index SNPs, we extracted corresponding summary statistics from the trans-ancestry meta-analysis for each phenotype. We computed the −log_10_(*P*-value) from the trans-ancestry meta-analysis, setting any trans-ancestry *P*>0.05 or with inconsistent directions of effect compared to the previous study equal to zero. From the resulting matrix of −log_10_(*P*-values), we generated a heatmap with R package pheatmap, and used hierarchical clustering to order the phenotype rows and the SNP columns in an unsupervised fashion.

### Discovery of Novel Phenotype-Locus Associations

Within the discovery EUR GWAS, we identified all loci that were suggestively associated (discovery EUR *P*<1×10^−5^) with any phenotype. For each of these suggestive associations, we designated the SNP with the lowest EUR *P*-value within a 500kb window the index SNP. From the resulting set of suggestive phenotype-locus association pairs, we determined whether the association replicated in one or more independent GWAS (replication EUR, LAT, or AA). We considered consistent direction of 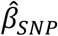 and replication population *P*<0.05 evidence of replication. Some of the index SNPs selected in the discovery EUR GWAS were not analyzed in the LAT and AA cohorts because the index SNP did not meet variant QC requirements (MAF>0.01 and Imputation R^2^>0.3) in one or both of those populations. For five of such phenotype-locus association pairs, there was another SNP that surpassed discovery EUR P<1×10^−5^ in the same region (<500Kb from the index SNP) *and* the alternative SNP was included in both non-EUR GWAS, so we used this alternative SNP to assess replication in the non-EUR cohorts. We also measured the Pearson correlation coefficient of 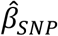 between the discovery EUR study and each of the three independent replication cohorts.

## Supporting information

Enlarged Figures

## Data Availability

This study replicates findings by large consortia, for which full summary statistics can be found at https://rgc-covid19.regeneron.com and https://www.covid19hg.org/results/.

https://www.ebi.ac.uk/ega/studies/EGAS00001004716

## AUTHOR CONTRIBUTIONS

GHLR, RP, and BR contributed equally to the manuscript. GHLR wrote the manuscript with substantial input from BR, RP, SCK, and DSP. DSP defined ancestry cohorts. RP and GHLR conducted all GWAS and meta-analyses with support from DSP. BR conducted literature review. MZ, DSP, DAT, SCK, MP, MG, LR, AHB, HG performed genotype imputation and data preparation. MVC and KAR designed the COVID-19 survey questionnaire and GHLR, SCK, MVC, KAR and SRM designed novel phenotypes. NB and MVC created the demographic table. ARG, AHB, and HG facilitated forward progression of the manuscript and provided input and guidance. The AncestryDNA Science Team contributed to additional work, allowing for the completion of the COVID-19 research and manuscript. KAR led the COVID-19 research and data teams. KAR, ELH, and CAB provided project guidance. All authors have contributed to and reviewed the final manuscript.

## AncestryDNA Science Team

Yambazi Banda, Ke Bi, Robert Burton, Marjan Champine, Ross Curtis, Karen Delgado, Abby Drokhlyansky, Ashley Elrick, Cat Foo, Jialiang Gu, Heather Harris, Shea King, Christine Maldonado, Evan McCartney-Melstad, Patty Miller, Keith Noto, Jingwen Pei, Jenna Petersen, Chodon Sass, Alisa Sedghifar, Andrey Smelter, Sarah South, Barry Starr, Cecily Vaughn, Yong Wang

## COMPETING INTERESTS

The authors declare competing financial interests: authors affiliated with AncestryDNA are employed by Ancestry and may have equity in Ancestry.

## ACKNOWLEDGEMENTS

We thank our AncestryDNA customers who made this study possible by voluntarily contributing information about their experience with COVID-19 through our survey. Without them, this work would not be possible. We additionally thank our collaborators at Regeneron Genetics Center and the COVID-19 Host Genetics Initiative for including us in ongoing meta-analyses aimed to improve understanding of COVID-19 infection susceptibility and severity.

## Notes

### Funding Statement

No external funding was received for this study.

### Author Declarations

All data for this research project were from subjects who provided prior informed consent to participate in AncestryDNAs Human Diversity Project, as reviewed and approved by our external institutional review board, Advarra (formerly Quorum). All data were de-identified prior to use.

