## Supplement for "Novel COVID-19 phenotype definitions reveal phenotypically distinct patterns of genetic association and protective effects"

**AncestryDNA Science Team:** Yambazi Banda, Ke Bi, Robert Burton, Marjan Champine, Ross Curtis, Karen Delgado, Abby Drokhlyansky, Ashley Elrick, Cat Foo, Jialiang Gu, Heather Harris, Shea King, Christine Maldonado, Evan McCartney-Melstad, Patty Miller, Keith Noto, Jingwen Pei, Jenna Petersen, Chodon Sass, Alisa Sedghifar, Andrey Smelter, Sarah South, Barry Starr, Cecily Vaughn, Yong Wang

### SUPPLEMENTARY FIGURES

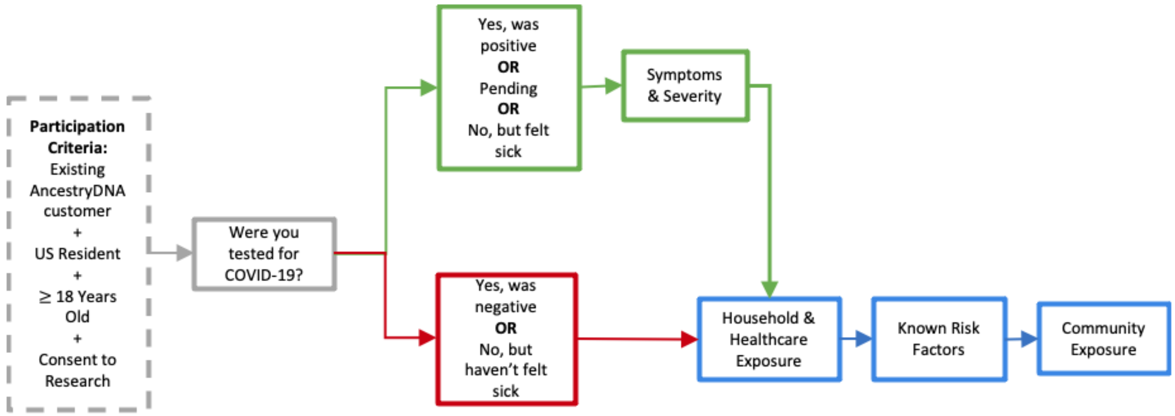


**Supplementary Figure 1. AncestryDNA COVID-19 Survey Flow.** Topical schema showing the flow and logic of the Ancestry COVID-19 survey. The gating question assesses whether the respondent has been tested for COVID-19 then shuttles respondents into two different flows: one with symptom, hospitalization and complication assessment, and one that does not include those questions. Answers that prompt the detailed sickness assessment are: positive COVID-19 nasal swab test, tested but with results pending, and not tested but have had flu-like symptoms with a fever at some point since the beginning of February 2020. All survey participants answer questions related to potential exposures and suspected risk factors for COVID-19 susceptibility and severity.

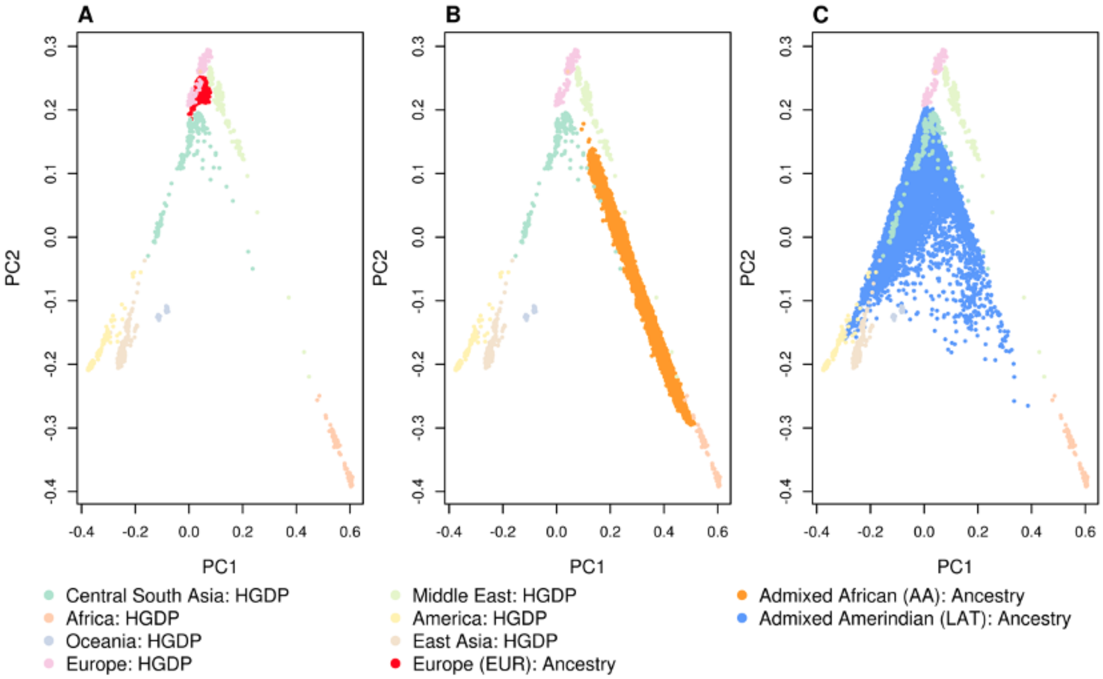


**Supplementary Figure 2. Principal Components Plot of the three genetic ancestry cohorts.** Principal component (PC) plots of the survey cohort together with samples from the Human Genetic Diversity Project superpopulations, denoted "HGDP". The PC1 versus PC2 plots are shown for the following cohorts: **(A)** European (EUR; Red), **(B)** Admixed African-European including 100% African ancestry (AA; Orange), and **(C)** Admixed Amerindian including 100% Amerindian ancestry (LAT; Blue).

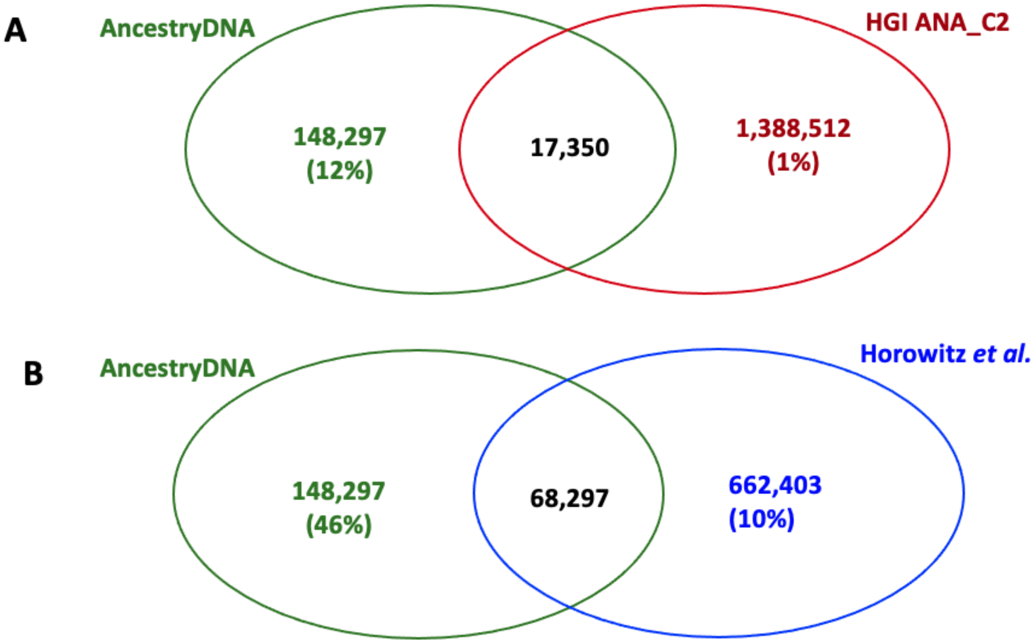


**Supplementary Figure 3. Sample overlap between AncestryDNA Study and (A) HGI, (B) Horowitz *et al.*** Venn diagrams show the number of samples (cases plus controls) that overlapped in COVID-19 GWAS meta-analyses. Green numbers represent the total number of subjects used in any of the eight phenotype GWAS described here, red numbers indicate the total number of subjects included in the largest HGI GWAS ("ANA_C2"), and blue numbers indicate the total number of subjects included in any study by Horowitz *et al*. Black numbers represent the sample overlap for each study. Percentages are the number of overlapping samples divided by the number of total subjects for each study.

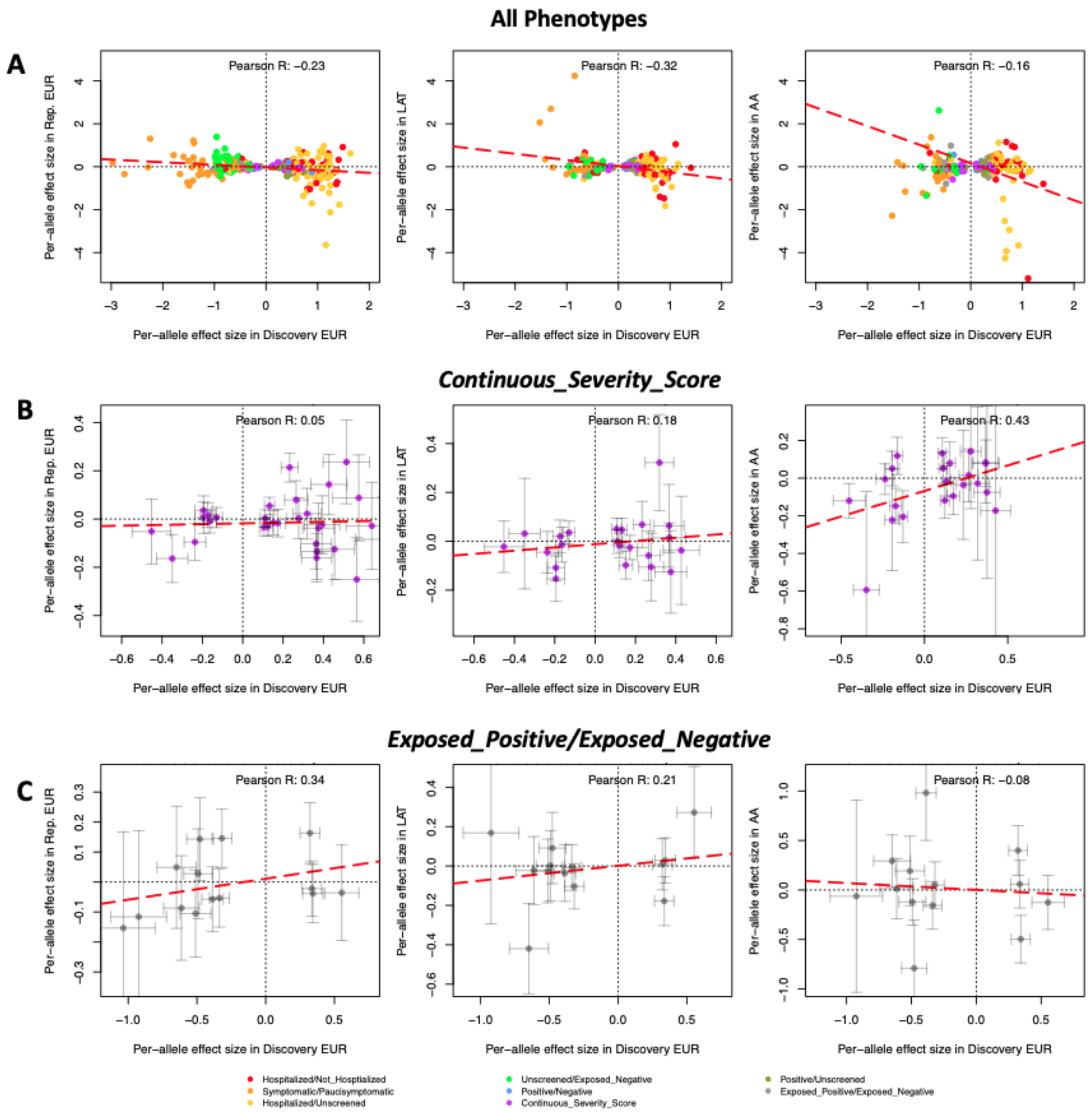


**Supplementary Figure 4. Correlation of effect estimates in the EUR discovery study with each of the three replication cohorts: replication EUR (left), LAT (middle), AA (right).** $\hat{\beta}_{SNP}$ comparison of 297 suggestive SNPs identified in the discovery EUR GWAS versus replication cohorts. In each plot, per-allele effect sizes for the minor allele in the discovery EUR GWAS is plotted along the x-axis, and the replication effect size is plotted on the y-axis in the replication EUR, LAT, and AA in the left, middle, and right plot, respectively. For the EUR replication cohort, all 297 SNPs are plotted, and for LAT and AA, a subset of 173 variants were plotted (see Methods). Each point represents the per-allele $\hat{\beta}_{SNP}$ and grey bars represent standard error. Points are colored by phenotype. Red dashed line denotes the line of best fit based on linear regression. **(a)** shows all suggestive discovered SNPs for all phenotypes, **(b)** suggestive discovered SNPs associated with the *Continuous_Severity_Score* phenotype, **(c)** suggestive discovered SNPs associated with the *Exposed_Positive/Exposed_Negative* phenotype.

### SUPPLEMENTARY TABLES

**Supplementary Table 1: AncestryDNA COVID-19 Survey Cohort Demographics**.

**Supplementary Table 2: GWAS Sample Sizes for Each of the Eight Phenotypes.**

**Supplementary Table 3: 13 Previously Identified COVID-19 Lead SNPs for Replication.** The minor allele in EUR is also the derived allele for all 13 SNPs.

**Supplementary Table 4: Association Results for 13 Previously Identified SNPs with the Eight Phenotypes.** Yellow highlighting indicates associations that replicated with a consistent direction of effect and trans-ancestry $P<0.05$.

**Supplementary Table 5: Review of Previously Reported *ABO* Associations with COVID-19 Susceptibility and Severity.**

**Supplementary Table 6: 297 Locus-Phenotype Association Pairs with Discovery EUR $P<1x10^{-5}$.** Results are available for all 297 associations in the two EUR studies, and a subset of 173 of these are available in the LAT and AA, and trans-ancestry meta-analysis studies. Blank entries indicate that the variant did not achieve QC thresholds for inclusion in that study.

**Supplementary Table 7: Summary of Independent Replication by Phenotype for the 297 Locus-Phenotype Association Pairs.**

**Supplementary Table 8: Extended Association and Replication Results for 15 Locus-Phenotype Association Pairs that Replicated in One or More Independent Populations.**
