## Supplementary material for "Novel COVID-19 phenotype definitions reveal phenotypically distinct patterns of genetic association and protective effects": Enlarged Figures

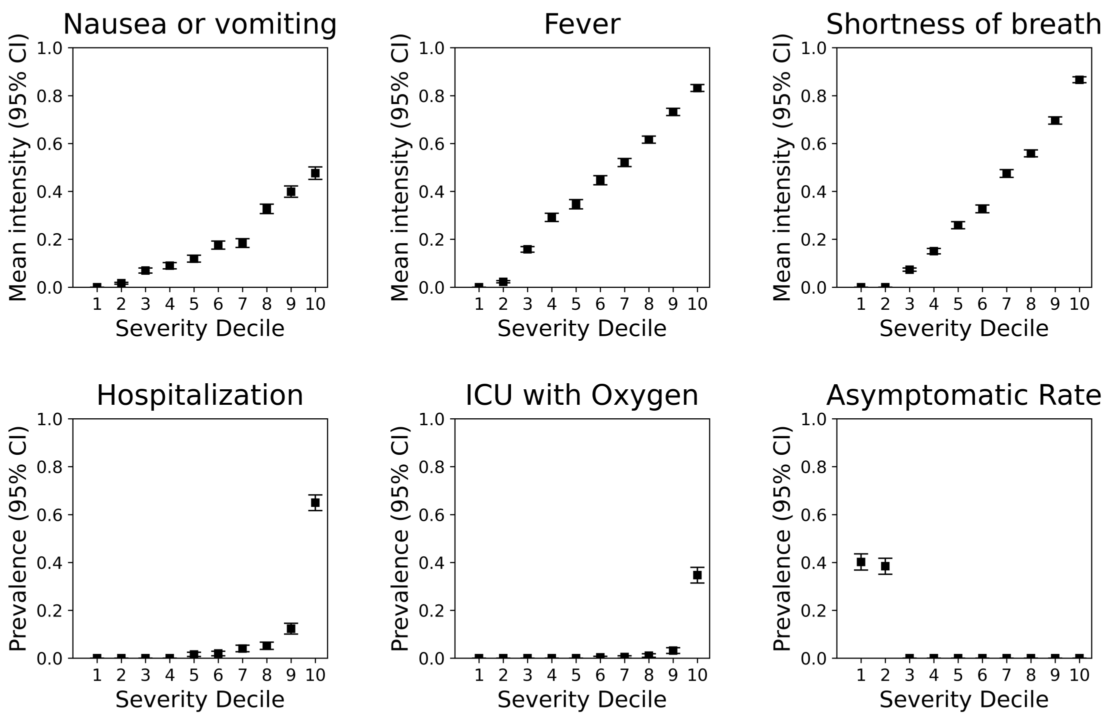


**Figure 1. COVID-19 Continuous Severity Score Captures Multiple Aspects of Symptom Severity Among COVID-19(+) Individuals.** The continuous severity score was derived from the first principal component across nine survey fields related to COVID-19 clinical outcomes, including three symptoms, hospitalization, ICU admittance, and other severe complications due to COVID-19 illness (see Methods). Plots reflect mean symptom severity (top three panels) or prevalence (bottom three panels) for several fields as a function of ascending severity decile. Symptom information was encoded as follows: 0=None, 0.2=Very Mild, 0.4=Mild, 0.6=Moderate, 0.8=Severe, and 1.0=Very Severe. A paucisymptomatic case corresponds to reporting symptoms of mild intensity or less. Squares represent the estimate and vertical lines represent the 95% confidence intervals for each estimate.

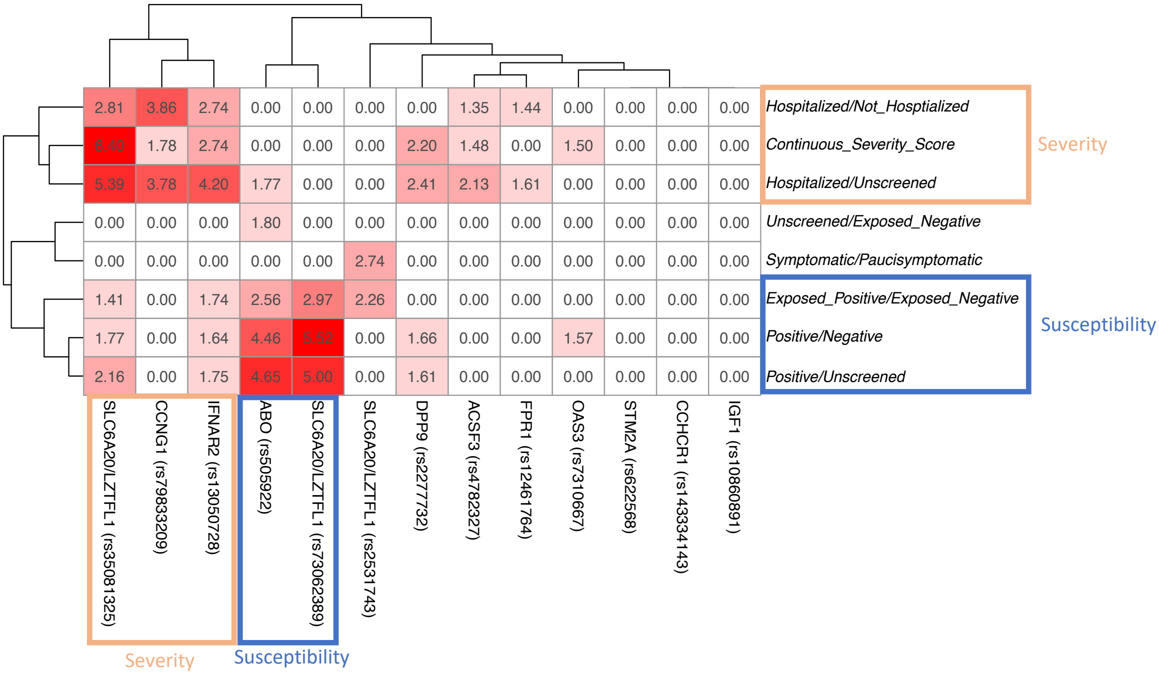


**Figure 2: Heatmap of replication at 13 lead SNPs identified by previous studies.** Each pairwise block represents the trans-ancestry meta-analysis -$\log_{10}$(*P*-value) for the association between one of the eight phenotypes we examined, and one of 13 loci previously identified by Horowitz *et al*. and/or HGI. Red blocks denote replication, with darker shades of red corresponding to lower trans-ancestry *P*-values in our analysis, and white blocks representing no association. All associations with trans-ancestry $P>0.05$ or with inconsistent directions of effect relative to the previous study were forced to have -$\log_{10}$(*P*-value)=0. SNP and phenotype labels were ordered by hierarchical clustering, with corresponding dendrograms shown on the top and left of the figure. Orange rectangles annotate phenotypes or loci that appear to associate more strongly with severity whereas blue rectangles annotate phenotypes or loci that appear to associate more strongly with susceptibility. Extended summary statistics for all associations in all studies are available in **Supplementary Table 4**.

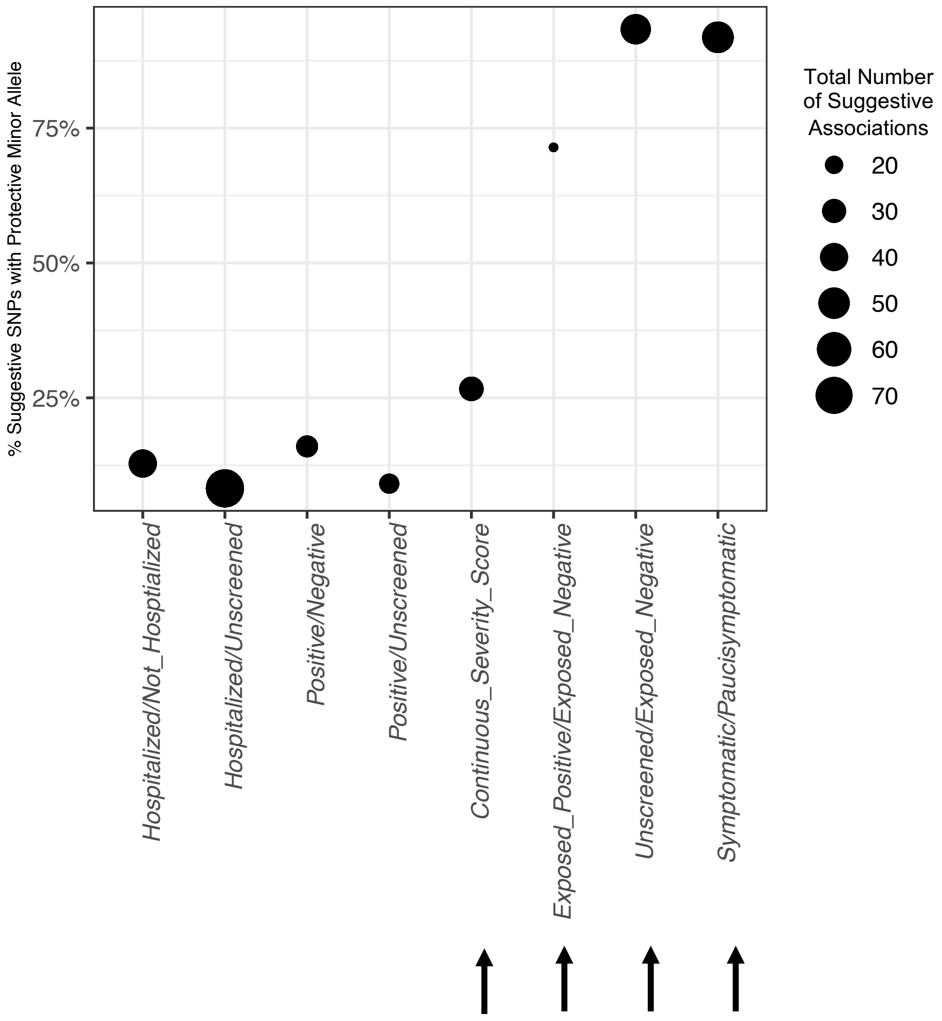


**Figure 3. Novel Phenotypes Detect More Associations with a Protective Minor Allele.** The size of each point represents the total number of novel, suggestive SNPs (discovery EUR $P<1x10^{-5}$) for each of the eight phenotypes. The y-axis position of each point shows the percentage of suggestively associated SNPs for which the discovery EUR minor allele was in the protective direction of effect. Arrows show the four novel phenotype definitions.

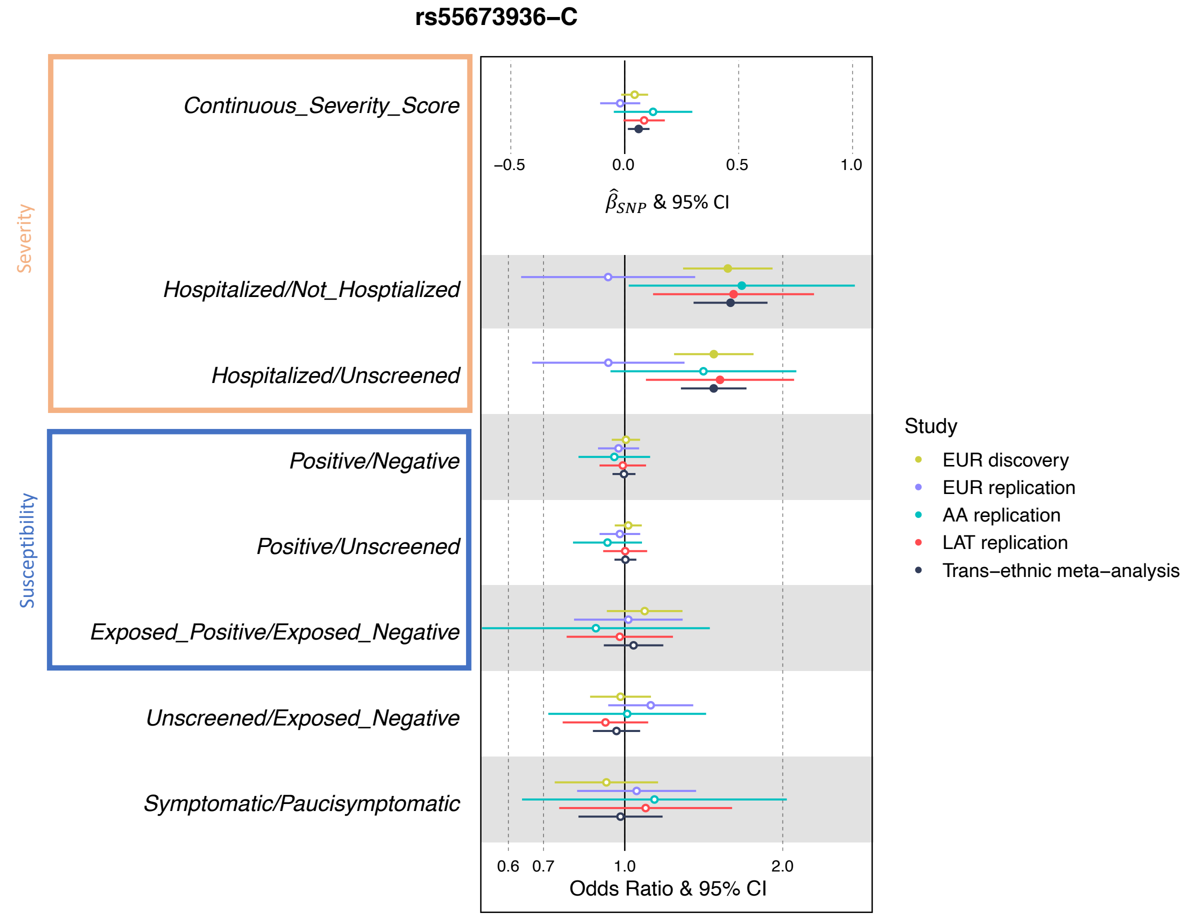


**Figure 4. Forest Plot of Novel Association with *GALNT18* intronic SNP, rs55673936-C, with the eight phenotypes**. Circles indicate effect estimates and horizontal lines represent 95% confidence intervals. *Continuous_Severity_Score* was the only continuous phenotype and therefore the reported effect estimate is the $\hat{\beta}_{SNP}$, which can be interpreted as severity score standard deviations from the mean per each copy of the "C" minor allele. For all other phenotypes, per-allele odds ratios are reported. Filled circles indicate $P$<0.05. The orange rectangle annotates phenotypes in the severity cluster and the blue rectangle annotates the susceptibility cluster, with clusters defined in **Figure 2**.
